# *AMROrbit*: Reimagining AMR Stewardship through an Open Source, Dynamic, Actionable Scorecard from Routine Microbiology Data

**DOI:** 10.1101/2025.06.05.25328985

**Authors:** Jasmine Kaur, Noel Abraham Tiju, Rishi Pendyala, Muthuraj Vairamuthu, Aryan Gupta, Tavpritesh Sethi

**Affiliations:** Center of Excellence in Healthcare, Indraprastha Institute of Information Technology Delhi, Okhla Industrial Estate Phase III, New Delhi, India, 110020; Department of Computational Biology, Indraprastha Institute of Information Technology Delhi, Okhla Industrial Estate Phase III, New Delhi, India, 110020

## Abstract

We present *AMROrbit* scorecard, an explainable model and an open-sourced tool leveraging routine microbiology data to enable data-driven AMR stewardship and surveillance. We built the scorecard on a global dataset of 229 antibiotic–microorganism–sample combinations collected across 84 countries using robust statistical models combined with an intuitive phase-space visualization of the amplitude and velocity of AMR trajectories. *AMROrbit* revealed rising resistance in 61.6% of combinations and spiraling-out trajectories in 18.1% of countries, signaling an urgent need for proactive data-driven stewardship in these areas.

## Main Text

There is a growing global recognition of antimicrobial resistance (AMR) as a silent pandemic^1–3^. To address this threat, effective stewardship and surveillance must be driven by early, actionable insights into shifting resistance patterns. However, the reliance on static antibiograms and fixed threshold alerts that only signal concern after resistance prevalence has already risen beyond safe limits^4–6^. By the time these conventional prompts arrive, clinicians and policymakers have often missed critical windows for updating empiric therapy guidelines, auditing antibiotic use or deploying rapid diagnostics, allowing resistance to spiral unchecked. Our previous work^7–8^ provided a proactive, model-based evaluation of resistance and its rate of change. However, the understanding of the dynamics of resistance was lacking.

To bridge this gap, we have developed *AMROrbit*, an open-source, intuitive, dynamic and explainable phase-space model for transforming stewardship-as-reporting to stewardship-as-strategy. By classifying trajectories as spiraling-in (towards Q1), spiraling-out (towards Q4), or persistent resistance, *AMROrbit* revealed significant global and country-specific resistance dynamics. To build this visualization, country-level resistance and its trend over time were estimated in rolling windows over a nine-year period. Using global medians as reference, each country’s estimates and their evolution were represented in four quadrants: AMR containment (spiral-in), persistent resistance, and emerging risk (spiral-out) (Figure 1a).

**Figure 1.**
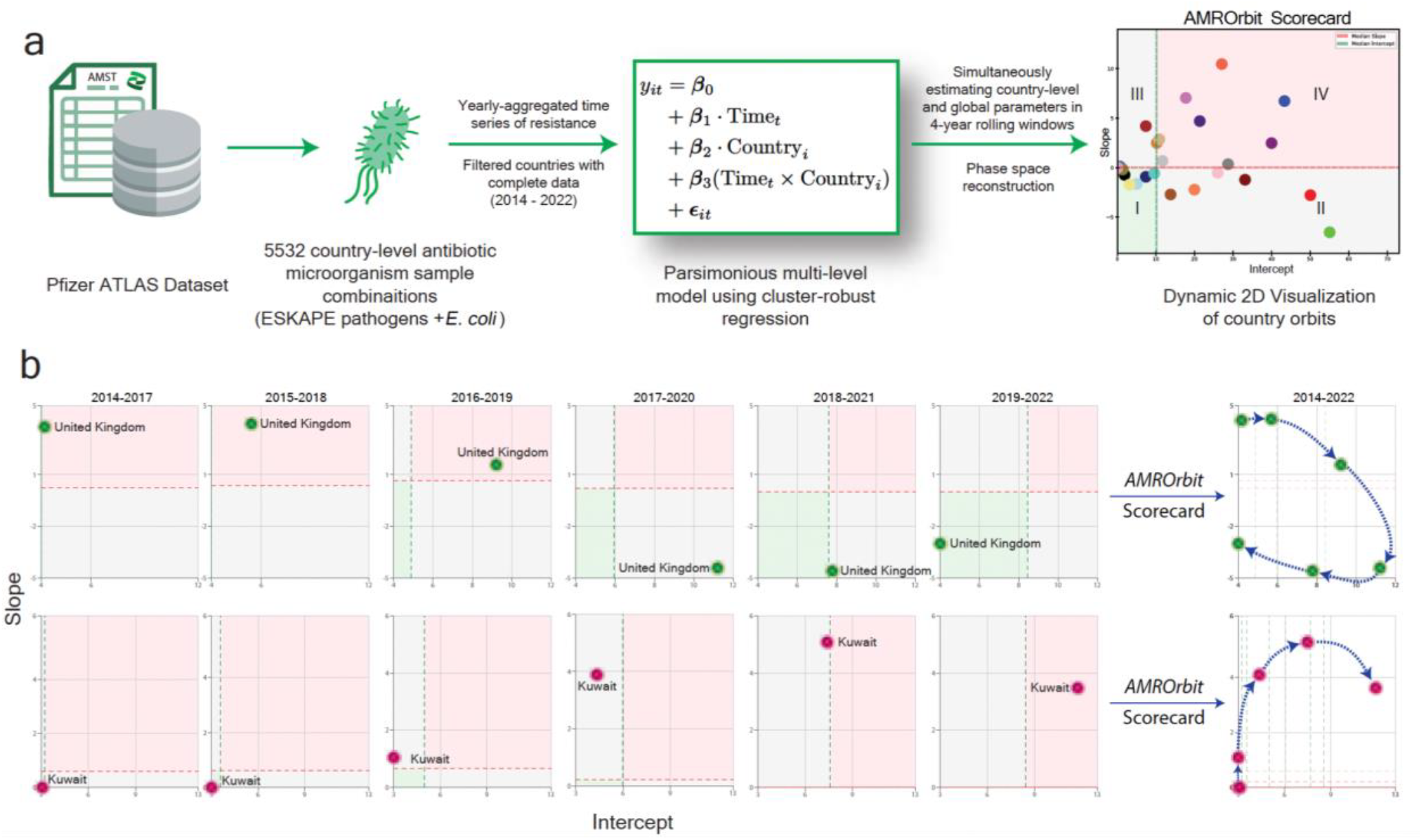
From Snapshots to Trajectories—AMROrbit pipeline and dynamic visualization of antimicrobial resistance (AMR) trajectories. a) Schematic representation of the *AMROrbit* scorecard and analytical pipeline. Cluster robust estimates of baseline and rate of change of resistance were computed from AMST data for ESKAPE pathogens and *E. coli* from 84 countries across multiple infection sources (urinary tract, bloodstream, sputum, and wound infections). Four-year rolling windows (with three-year overlap) to simultaneously estimate global and country-specific AMR rates were visualized as a dynamic phase-space reconstruction to reveal country-specific trajectories. b) *AMROrbit* Scorecard. Country trajectories are classified into intuitive patterns: spiraling-in (positive AMR containment), persistent resistance (remaining in the same quadrant), and spiraling-out (negative AMR trajectory). The United Kingdom demonstrates a positive trajectory, progressively moving towards reduced resistance levels. Conversely, Kuwait’s trajectory signals deterioration, shifting from Quadrant I to IV, indicating an urgent need for targeted intervention.

To assess the utility of *AMROrbit*, we demonstrate its application to ESKAPE pathogens and *E. coli* using a global dataset comprising samples from 84 countries collected between 2004 and 2022. This dataset was sourced from the Pfizer ATLAS repository as part of the Vivli AMR Surveillance Data Challenge^9–11^. We analyzed 5,532 country-level antibiotic–microorganism–sample combinations with complete annual data spanning 2014 to 2022, enabling longitudinal assessment of resistance trends across consistent observational units.

For a demonstrative example (Figure 1b), Kuwait’s *K*.*pnemoniae-*Amikacin*-*blood trajectory for 2014-2017 remained on the global median with decreasing slope (−0.18). However, from 2015-2018, the slope increased and consequently, in the next subsets we observed a spiralling-out trajectory into the high slope-high intercept quadrant. In the 2015-2018 subset, the model detected a rising slope that, by the 2019-2022 subset, pushed the country into the critical-escalation quadrant two subsets before its resistance rate and baseline increased farthest away from the global medians. Thus, *AMROrbit* provides actionable early-warning signals that can prompt intervention long before resistance rates reach critical levels.

Further, aggregated global results revealed 61.6% of antibiotic–microorganism–sample combinations showed rising baseline resistance. Among country-level combinations, 18.1% were spiralling-out (e.g. Kuwait in Figure 1b), whereas only 14.2% were spiralling-in (e.g. the United Kingdom in Figure 1b), suggesting opportunities for policy learning and adaptation (Table 1). These quadrant visualizations offer quantitative benchmarks for cross-country comparisons and can inform the allocation of global stewardship resources to the highest-risk settings. Organism-specific patterns further suggest where strategic interventions could yield maximum benefit. *Enterobacter species* in sputum samples exhibited the highest spiraling-out proportions, followed by *A. baumanii* in sputum, *A. baumanii* in urine, *P. aeruginosa* in sputum and *E. coli* in urine.

**Table 1.**
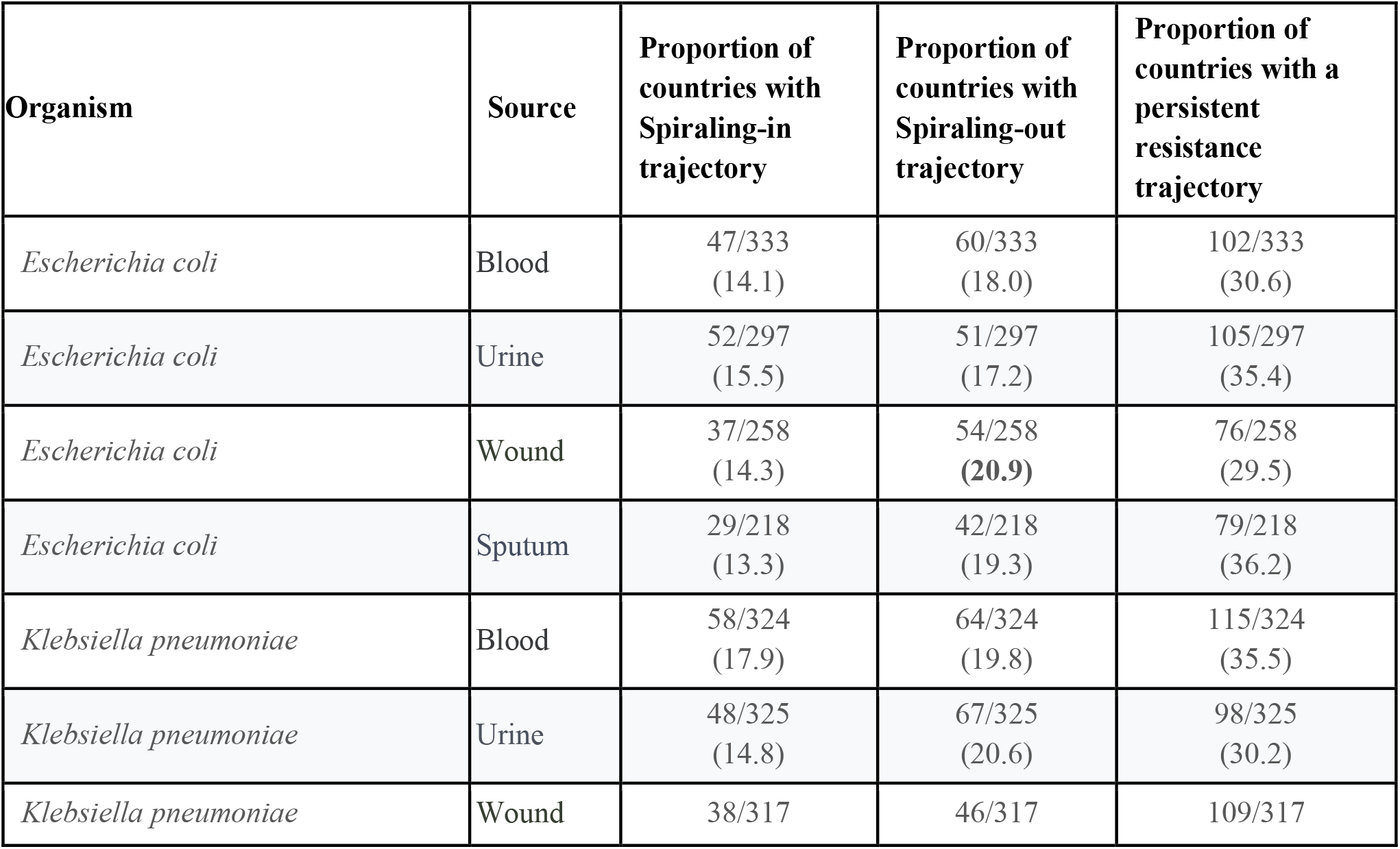
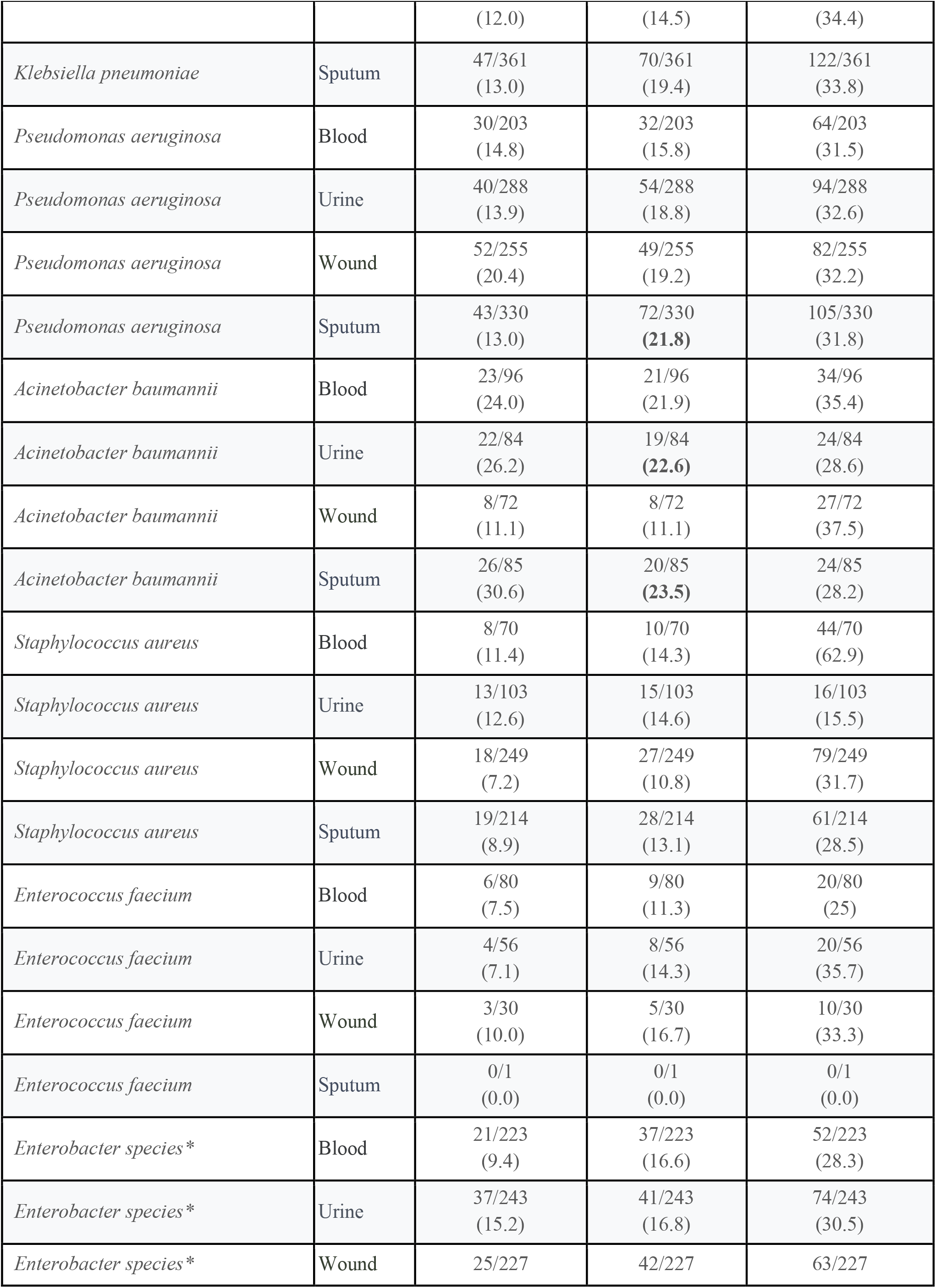
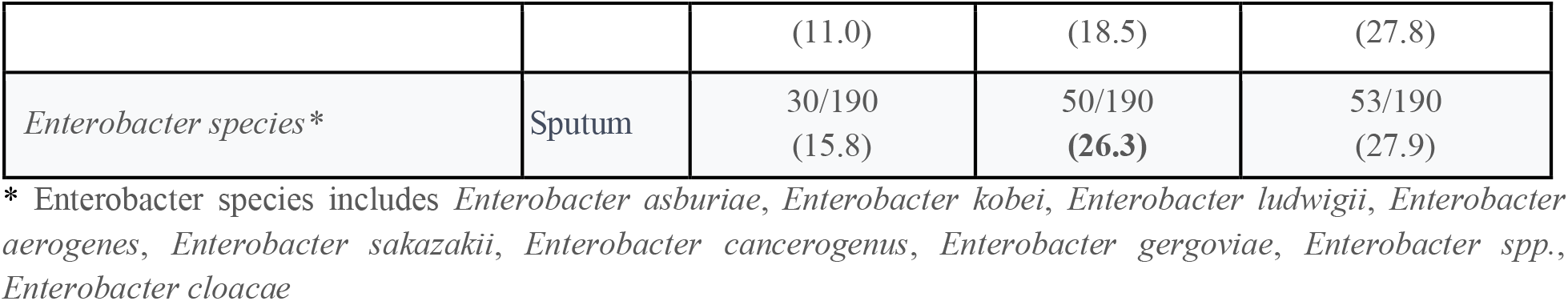
The proportion of countries depicting spiralling-in, spiralling-out, and persistent resistance trajectories for 5,532 country-level antibiotic-microorganism-sample combinations. (Spiraling-in trajectory is calculated as movement from any quadrants (II, III or IV) to I; Spiraling-out trajectory is calculated as movement from any quadrants (I, II or III) to IV; persistent resistance trajectory for countries remaining in their quadrants)

These results underscore the need for systems that move beyond static prevalence measures to characterize the dynamic, temporal nature of AMR. *AMROrbit* fulfils this need by not only quantifying resistance trajectories but also contextualizing them within a policy-relevant quadrant framework. In doing so, it reveals opportunities for early intervention, cross-country benchmarking, and adaptive policy design.

*AMROrbit’s* transparent, regression-based flags integrate seamlessly with existing WHO GLASS estimation principles, ensuring compatibility with global surveillance frameworks. Unlike black-box models, *AMROrbit* provides interpretable scorecards enabling discussions among analysts, clinicians, and policymakers. Its open-source GitHub repository available at https://github.com/tavlab-iiitd/AMROrbit and real-time dashboard (available at https://amrorbit.tavlab.iiitd.edu.in:3002) enables interactive exploration of trends, empowering stakeholders to track resistance shifts, compare trajectories, and design timely, data-driven responses

Our approach has some limitations. First, the reliance on complete yearly data may exclude countries or organisms with limited reporting, potentially affecting the detection of resistance trends. Second, the method assumes linear patterns within rolling time windows, which may not fully capture more complex or nonlinear changes. Although currently shown for the Pfizer ATLAS dataset, the framework is adaptable to other AMR datasets and supports bespoke levels of geographic and temporal granularity for dynamic trajectory monitoring and is currently under validation at two large tertiary care settings in New Delhi, India. Finally, while *AMROrbit* supports AMR stewardship, causal inference requires integration with additional contextual data (e.g., antibiotic consumption, policy interventions, or healthcare infrastructure).

Our open-source repository and interactive web application allow contextualization, transparent testing and extension for local settings. The tool is platform-independent, user-friendly, and adaptable to any AMST dataset in a standard format. By transforming retrospective surveillance data into dynamic and explainable scorecards, *AMROrbit* offers a scalable and interpretable framework that is both globally aligned and locally adaptive, ensuring timely and context-specific AMR stewardship.

## Data Availability

This analysis is based on research using data from Pfizer, obtained through https://amr.vivli.org.

## Acknowledgement

This analysis is based on research using data from Pfizer, obtained through https://amr.vivli.org. This work was supported by The Trinity Challenge, Indian Council of Medical Research (BMI/12(92)/2021) and Grand Challenges India (BT/GCI-AI0805/10/23).

## Code availability

All custom scripts and code for *AMROrbit* scorecard presented in this study are publicly available on the GitHub repository at https://github.com/tavlab-iiitd/AMROrbit. The real-time dashboard and a web-based application to build custom scorecards are available at https://amrorbit.tavlab.iiitd.edu.in:3002.

## Author Contributions

TS and JK conceptualised the study and designed the analytical framework. JK, NT, RP, MV, and AG performed the data analysis. TS and JK drafted the manuscript and prepared all figures. All authors have critically reviewed the manuscript and approved the final version.

## Online Methods

Yearly aggregated resistance percentages as described in previous studies^7,8^ were calculated for each antibiotic-organism-sample-country combination to generate a time series of resistance as

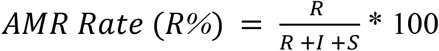

where R is the count of resistant isolates, S is the count of susceptible isolates, I is the count of Intermediate isolates

Countries with complete time series (aggregated values for all years from 2014 to 2022) for the entire duration were used for modelling. A hierarchical multi-level regression model was developed with resistance percentage as a dependent variable varying as a function of time, country, and their interaction effects. The model is designed to capture both main effects (time and country) and their interactions, allowing the relationship between resistance trends and time to vary by country.

The regression equation is:

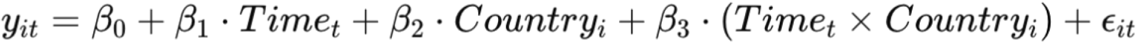

Where:

- y_it_ = AMR resistance percentage for country i at time t
- Time_t_ = Temporal variable (e.g., year in this case)
- Country_i_ = Categorical variable for the country
- β_0_,β_1_,β_2_,β_3_ = Model coefficients (intercept, slopes, and interaction term)
- ϵ_it_ = Error term

This model ensures that each country can have its own slope, reflecting different AMR trends over time. The interaction term (Time_t_ × Country_i_) allows for country-specific trends, addressing heterogeneity across countries.

### Cluster-Robust Adjustments

To adjust for the clustering of data corresponding to hospital sites, cluster-robust sandwich estimates of the slope (trend) of resistance with its standard error and p-values were obtained using the sandwich packages. To account for heteroskedasticity (non-constant variance) and intra-cluster correlations (dependence within countries), the standard errors were adjusted using the “sandwich” estimator, given by:

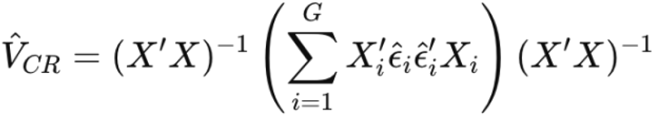

Where:

- X = Design matrix
- ϵ^i = Residuals for cluster i (e.g., for each country)
- G = Number of clusters (countries)

This ensures the model provides reliable parameter estimates with accurate standard errors and p-values.

### Phase-Space Representation to derive *AMROrbit scorecard*

The country-level and global parameters were simultaneously modelled in four-year subsets with three year overlap for each combination. For the scorecard, the slopes and the intercepts corresponding to each country were extracted and represented as baseline resistance (X-axis) and rate of change in resistance (Y-axis). The quadrants, derived using global medians, were as follows: a) Quadrant I with low baseline resistance and low slope, b) Quadrant II with low slope but high baseline resistance, c) Quadrant III with high slope but low baseline resistance d) Quadrant IV with high slope and high baseline resistance. Scorecards developed and visualized across subsets allowed us to visualize country trajectories in terms of resistance and how quickly it is changing.

## References

1. Antimicrobial resistance: a silent pandemic. Nat. Commun. 15, 6198 (2024). 10.1038/s41467-024-50457-z

2. World Health Organization. Global Action Plan on Antimicrobial Resistance. WHO, Geneva (2015).

3. Laxminarayan R. The overlooked pandemic of antimicrobial resistance. Lancet 399, 606–607 (2022).

4. Truong WR, Hidayat L, Bolaris MA, Nguyen L, Yamaki J. The antibiogram: key considerations for its development and utilization. JAC Antimicrob. Resist. 3, dlab060 (2021).

5. Hindler JF, Stelling J. Analysis and presentation of cumulative antibiograms: a new consensus guideline from the Clinical and Laboratory Standards Institute. Clin. Infect. Dis. 44, 867–873 (2007).

6. Fu Q, Zhang Y, Shu Y, Ding M, Yao L, Wang C. From data to action: charting a data-driven path to combat antimicrobial resistance. arXiv preprint arXiv:2502.00061 (2025)

7. Kaur J, Singh H, Sethi T. Emerging trends in antimicrobial resistance in bloodstream infections: a multicentric longitudinal study in India (2017-2022). Lancet Reg. Health Southeast Asia 10, 100132 (2024).

8. Kaur J, et al. SSRN [Preprint]. https://ssrn.com/abstract=4316853 (2023).

9. Pfizer. The ATLAS database: A global surveillance tool for antimicrobial resistance. https://amr.vivli.org (accessed 2 Apr 2025).

10. Vivli AMR. Pfizer – ATLAS. https://amr.vivli.org/faq/atlas/ (2022, accessed 2 Apr 2025)

11. Vivli AMR. 2023 Data Challenge Overview. https://amr.vivli.org/data-challenge/data-challenge-overview-2023/ (accessed Apr 2025).

